# Longitudinal effects of CPAP therapy on MRI-Perivascular spaces in obstructive sleep apnoea

**DOI:** 10.64898/2026.03.16.26348478

**Authors:** William Pham, Donggyu Rim, Alexander Jarema, Zhibin Chen, Mohamed Salah Khlif, Amy Brodtmann, Luke A. Henderson, Vaughan G. Macefield

**Author notes:** Corresponding author: Professor Vaughan G. Macefield, Monash University, School of Translational Medicine Department of Neuroscience, Level 6, 99 Commercial Road, Melbourne, VIC, 3004.

## Abstract

Obstructive sleep apnoea (OSA) is a disorder marked by repeated episodes of airway collapse during sleep, leading to hypoxaemia and sympathoexcitation. Its impact on brain fluid transport remains unclear. We investigated MRI-visible perivascular spaces (PVS) in healthy controls (*n*=20; 5 females, mean±SD age = 52.1±9.9 years) and OSA patients (*n*=20; 3 females, mean±SD age = 54.6±9.6 years) before and after continuous positive airway pressure (CPAP) therapy in a longitudinal case-control study. MRI-PVS were automatically quantified using a deep learning model called the nnU-Net. At baseline, OSA patients had significantly greater whole-brain PVS volumes and cluster counts than controls (volume: exp(β)=1.65, 95%CI [1.07, 2.51], *p*=0.01; cluster counts: exp(β)=1.51, 95%CI [1.1, 2.04], *p*=0.01). However, after 12 months of CPAP, these differences were no longer significant (volume: exp(β)=1.56, 95%CI [1.03, 2.39], *p*=0.054; cluster counts: exp(β)=1.39, 95%CI [0.97, 1.92], *p*=0.072). Similarly, PVS metrics were significantly greater in OSA patients than controls at baseline in the frontal (volume: exp(β)=1.66, 95%CI [1.02, 2.64], *p*=0.04; cluster counts: exp(β)=1.46, 95%CI [1.02, 2.08], *p*=0.04) and temporal lobe (volume: exp(β)=1.92, 95%CI [1.2, 3.03], *p*=0.01; cluster counts: exp(β)=1.68, 95%CI [1.1, 2.55], *p*=0.02). After 12 months of CPAP, PVS metrics remained significantly higher in the OSA patients compared to controls in the frontal (volume: exp(β)=1.68, 95%CI [0.94, 2.87], *p*=0.085; cluster counts: exp(β)=1.4, 95%CI [0.92, 2.05], *p*=0.12) and temporal lobes (volume: exp(β)=1.54, 95%CI [0.94, 2.38], *p*=0.085; cluster counts: exp(β)=1.44, 95%CI [0.95, 2.12], *p*=0.089). These findings suggest that OSA is associated with PVS enlargement, which may be regionally reversible with CPAP treatment.

## Introduction

Obstructive sleep apnoea (OSA) is a prevalent sleep disorder characterised by recurrent episodes of upper airway obstruction during sleep. The ensuing episodes of intermittent hypoxia and fragmented sleep architecture leads to a range of cognitive and behavioural symptoms, including excessive daytime sleepiness, impaired concentration, mood disturbances and emotional dysregulation (Lévy et al., 2015). In addition, the long-lasting effects of nocturnal hypoxaemia lead to increases in muscle sympathetic nerve activity (MSNA) during the day, resulting in elevated blood pressure (Carlson et al., 1993; Fatouleh et al., 2014, 2015; Hedner et al., 1988; Henderson & Macefield, 2016; Lundblad et al., 2014; Somers et al., 1995). OSA frequently co-occurs with systemic conditions such as obesity and metabolic syndrome, further compounding its impact on long-term health (Lévy et al., 2015). Furthermore, magnetic resonance imaging (MRI) studies have shown that OSA is associated with significant changes in brain structure and function (Fatouleh et al., 2014, 2015). Given these widespread negative effects, there is a critical need to better understand the neurobiological consequences of OSA, particularly in relation to brain health and fluid homeostasis.

A key driver of fluid homeostasis and waste clearance in the brain is the glymphatic system. It operates through interconnected channels of perivascular spaces to circulate the movement of interstitial fluid and solutes. Glymphatic activity is most pronounced during deep, slow-wave sleep (Xie et al., 2013). Disruptions to sleep are known to impair glymphatic clearance, leading to the accumulation of neurotoxic proteins such as amyloid-β (Xie et al., 2013). Experimental models have shown glymphatic impairment accelerates cognitive decline and is associated with increased amyloid-β deposition (Deng et al., 2024; Xie et al., 2013).

Perivascular spaces (PVS), which are integral to glymphatic flow, are detectable on MRI and considered an emerging biomarker of cerebral small vessel disease (SVD) (Duering et al., 2023). Several studies have reported an increased burden of MRI-visible PVS in individuals with OSA (Huang et al., 2019; Jia et al., 2021). Additionally, meta-analytic evidence links OSA to several neuroimaging markers of SVD including PVS (G. Lee et al., 2023). Moreover, as noted above, OSA is linked to elevated sympathetic vasoconstrictor drive, which may further impede cerebrovascular dynamics (Henderson & Macefield, 2016). MSNA-coupled fMRI has revealed significant changes in Blood Oxygen Level Dependent (BOLD) signal intensity in several cortical and subcortical sites in OSA (Fatouleh et al., 2014; Lundblad et al., 2014), including sites within the key brainstem region containing sympathetic premotor neurons - the rostral ventrolateral medulla (RVLM) – that are normalised following effective continuous positive airway pressure (CPAP) therapy, which also reduces MSNA (Fatouleh et al., 2015; Lundblad et al., 2015).

Given the potential association between changes in PVS measurements and elevated sympathetic vasoconstrictor drive in OSA, we applied a deep learning model, nnU-Net, to investigate PVS changes in a case-control cohort of OSA patients and healthy controls. We hypothesised that OSA patients would exhibit increased PVS burden at baseline (prior to CPAP therapy) compared to controls, reflecting disturbed cerebrovascular dynamics. In addition, we hypothesised that, given structural and functional changes seen in OSA (Fatouleh et al., 2015; Lundblad et al., 2015), PVS burden would be reduced by 6 and 12 months of CPAP therapy and that these reductions would correlate with the reductions in MSNA following treatment.

## Methods

### Subjects

A total of 40 participants were included in the study, comprising 20 healthy controls and 20 patients with OSA. All OSA patients underwent overnight in-laboratory polysomnography to confirm diagnosis. Daytime sleepiness was assessed in all participants using the Epworth Sleepiness Scale questionnaire. Healthy controls underwent overnight home-based apnoea screening using a ResMed ApneaLink^TM^ device, which continuously monitored nasal airflow and peripheral oxygen saturation (SaO_2_).

The study was approved by the Human Research Ethics Committees of the University of New South Wales and Prince of Wales Hospital. All participants provided informed written consent in accordance with the Declaration of Helsinki.

### Physiological recordings

Physiological measurements were obtained prior to MRI scanning. Mean systolic and diastolic blood pressure (SBP/DBP), heart rate (HR), and respiratory rate were recorded. Each parameter was continuously monitored for 10 minutes while participants were at rest, with final values derived from the mean of the last 5 minutes of recording. Continuous blood pressure measurements were acquired via radial arterial tonometry (Colin 7000 NIBP; Colin Corp., Aichi, Japan). Electrocardiographic activity was recorded using Ag-AgCl surface electrodes in a three-lead configuration, on the chest, and respiration was measured using a strain-gauge transducer around the chest (Pneumotrace; UFI, Morro Bay, CA, USA).

### Muscle Sympathetic Nerve Activity Recording

MSNA was recorded using a tungsten microelectrode percutaneously inserted into the right common peroneal nerve. Nerve signals were amplified (gain 2 × 10^4^; bandpass 0.3–5.0 kHz) using a low-noise, electrically isolated head-stage (NeuroAmpEX, ADInstruments, Australia). The recorded signal was root-mean-square (RMS) processed with a moving average window of 200 ms. A digital high-pass filter (300 Hz) was applied to further reduce scanner noise. Bursts of MSNA were manually counted from the RMS-processed signal, and burst frequency (bursts per minute) and burst incidence (bursts per 100 heartbeats) quantified. The detailed procedures for MSNA recording were implemented as previously described (Fatouleh et al., 2014; Lundblad et al., 2014).

### MRI acquisition

High-resolution T1-weighted (T1w) MRI brain scans were acquired using a 3 Tesla Philips Achieva scanner equipped with a 32-channel SENSE head coil. Imaging was performed with a magnetisation-prepared repaid gradient echo (MPRAGE) sequence (turbo field echo; echo time=2.5 ms, repetition time=5600 ms, flip angle=8°, voxel size=0.8 mm^3^, 200 sagittal slices).

### MRI preprocessing

Perivascular spaces were automatically segmented from T1w MRI scans using a deep learning model based on the nnU-Net residual encoder architecture, as previously described (Isensee et al., 2021; Pham et al., 2025). The model generates separate labels for PVS within the white matter and basal ganglia, enabling direct quantification of PVS volumes and cluster counts in each region. Individual PVS clusters were identified with the ‘measure.label’ function from the ‘sci-kit image’ package in Python (v3.9.13). In addition, specialised nnU-Net models were used to segment PVS in the midbrain and hippocampi, with PVS volumes and cluster counts quantified similarly. All PVS segmentations were performed in the native anatomical space of each image. nnU-Net inference was conducted with an Nvidia A40 GPU on the MASSIVE high-performance computing cluster (Goscinski et al., 2014).

PVS measurements were quantified according to two brain atlases. The ICBM2009c asymmetric atlas was used to parcellate the white matter into frontal, parietal, temporal, and occipital lobes (Fonov et al., 2009). A vascular territory atlas was used to segment brain tissue according to the anterior cerebral artery (ACA), middle cerebral artery (MCA), and posterior cerebral artery (PCA) territories (Liu et al., 2023).

Brain extraction was performed using FastSurfer (v2) (Henschel et al., 2020). Using the Advanced Normalization Tools (ANTs, v2.4.3) (Avants et al., 2011), brain volumes were spatially registered to the MNI152 template. Rigid, affine and symmetric diffeomorphic normalisation was performed with antsRegistrationSyn.sh to produce forward and inverse transformations. Subject-specific atlases were generated by applying the inverse warp fields and affine transformations to both atlases, enabling native-space parcellation of PVS according to lobar and vascular territories.

We performed a validation procedure to assess the reliability of PVS measurements. Model predicted PVS cluster counts were compared against manual PVS counts. In the centrum semiovale (CS) and basal ganglia (BG), PVS were manually counted in two representative axial slices. Model predicted PVS counts from the same axial slices were compared to the manual PVS counts in the respective axial slices. Additionally, all PVS were manually counted in the midbrain and hippocampal regions. Model predicted PVS counts in the midbrain and hippocampal regions were compared to the manual PVS counts in these regions. Manual PVS counting of all baseline MRI scans was performed by a single experienced rater (WP) and independently reviewed by a board-certified radiologist (AJ).

### Statistical analysis

We used Lin’s concordance correlation coefficient (CCC) to assess the agreement between automated and manual PVS counts. Spearman’s rank correlation coefficient was used to assess the association between automated and manual PVS counts. The validation procedure was performed for PVS measurements in the CS, BG, midbrain, and hippocampus.

Next, we constructed generalised linear models (GLMs) to assess baseline group differences in PVS measurements. Normality of baseline PVS distributions was evaluated using Shapiro-Wilk tests; all metrics deviated from a normality (*p* < 0.05). Accordingly, GLMs with a Tweedie distribution and log-link function were used. All models were adjusted for age, sex, and total brain volume (TBV), which are established determinants of MRI-visible perivascular space measurements (Evans et al., 2022; Francis et al., 2019; Lynch et al., 2023).

TBV was derived from T1w MRI scans using FastSurfer (v2) and included as a covariate in the GLMs (Henschel et al., 2020). However, due to the limited sample size, several models failed to converge. Therefore, TBV was therefore excluded from models assessing PVS in the midbrain and hippocampus.

Separate GLMs were built for each PVS metrics, with the metric as the dependent variable and fixed effects for diagnosis, age, sex, and TBV. To ensure model validity, residual normality and heteroscedasticity were visually inspected, and covariates were pruned as needed to meet model assumptions. Robust 95% confidence intervals for all fixed effects were estimated using parametric bootstrapping with 1,000 replicates.

Since PVS data for controls were only available at baseline, additional GLMs were constructed to compare controls at baseline with OSA participants after 6 and 12 months of CPAP therapy. These models retained the same structure, but PVS values for OSA participants were substituted with measurements at 6 or 12 months, respectively.

To examine associations between physiological parameters (HR, SBP, DBP, MSNA burst frequency, and burst incidence) and PVS metrics, generalised linear mixed-effects models with a Tweedie distribution and log-link function were fitted. Each model included a PVS metric as the dependent variable and a physiological variable as an independent predictor, with adjustment for baseline age and timepoint. A random intercept was included for each subject. Covariates were systematically pruned to facilitate model convergence and ensure adherence to model assumptions. Statistical significance was defined as *p* < 0.05. Results from all models were adjusted for multiple comparisons with the Benjamini-Hochberg false discovery rate (FDR) correction.

Statistical analyses were conducted in R (v4.2.1). Concordance correlation coefficients were computed using the *EpiR* package. Generalised linear models were fitted using the *glmmTMB* package.

## Results

A total of 40 participants (8 females, mean±SD age = 53.4±9.7 years), comprising 20 healthy controls (5 females, mean age=52.1±9.9 years) and 20 OSA patients (3 females, mean age=54.6±9.6 years) were included in the baseline dataset (Table 1). Across the entire cohort, the median [Q1, Q3] PVS volume was 1,433.1 mm^3^ [988.3, 2,065.2], and the median [Q1, Q3] PVS cluster count was 165 [123.5, 267.5]. Baseline descriptive statistics of all PVS metrics are shown in Supplementary Table 1. Of the 20 OSA patients, 16 returned for the 6-month follow-up, and 15 returned for the 12-month follow-up (Table 2). However, four MRI scans from OSA subjects were excluded due to poor image quality, resulting in 14 usable scans at the 6-month follow-up and 13 at the 12-month follow-up.

**Table 1.** Demographic characteristics of the cohort at baseline stratified by diagnosis. Numerical variables are presented as mean±SD or median [Q1, Q3] for normally or non-normally distributed variables, respectively. Group differences in categorical variables were compared with Chi-squared tests. Group differences in numerical variables were compared with independent samples t-tests.

|  | **Controls**<br>**(*n* = 20)** | **OSA**<br>**(*n* = 20)** | **Total**<br>**(*N* = 40)** | ***p*-value** |
| --- | --- | --- | --- | --- |
| **Age, years** | 52.1±9.9 | 54.6±9.6 | 53.4±9.7 | 0.43 |
| **Sex, *n* female (%)** | 5 (25%) | 3 (15%) | 8 (20%) | 0.43 |
| **PVS volume (mm<sup>3</sup>)** | 1,195.1 [945.3, 1,493.4] | 1,752.3 [1035.5, 3,604.1] | 1,433.1 [988.3, 2,065.2] | 0.03 |
| **PVS cluster count** | 155.5 [119.3, 205.3] | 247.5 [140.8, 380.8] | 165 [123.5, 267.5] | 0.02 |
| **Session** |  |  |  |  |
| Baseline, *n* | 20 | 20 | 40 |  |
| 6 Month, *n* | – | 14 | – |  |
| 12 Month, *n* | – | 13 | – |  |

**Table 2.**
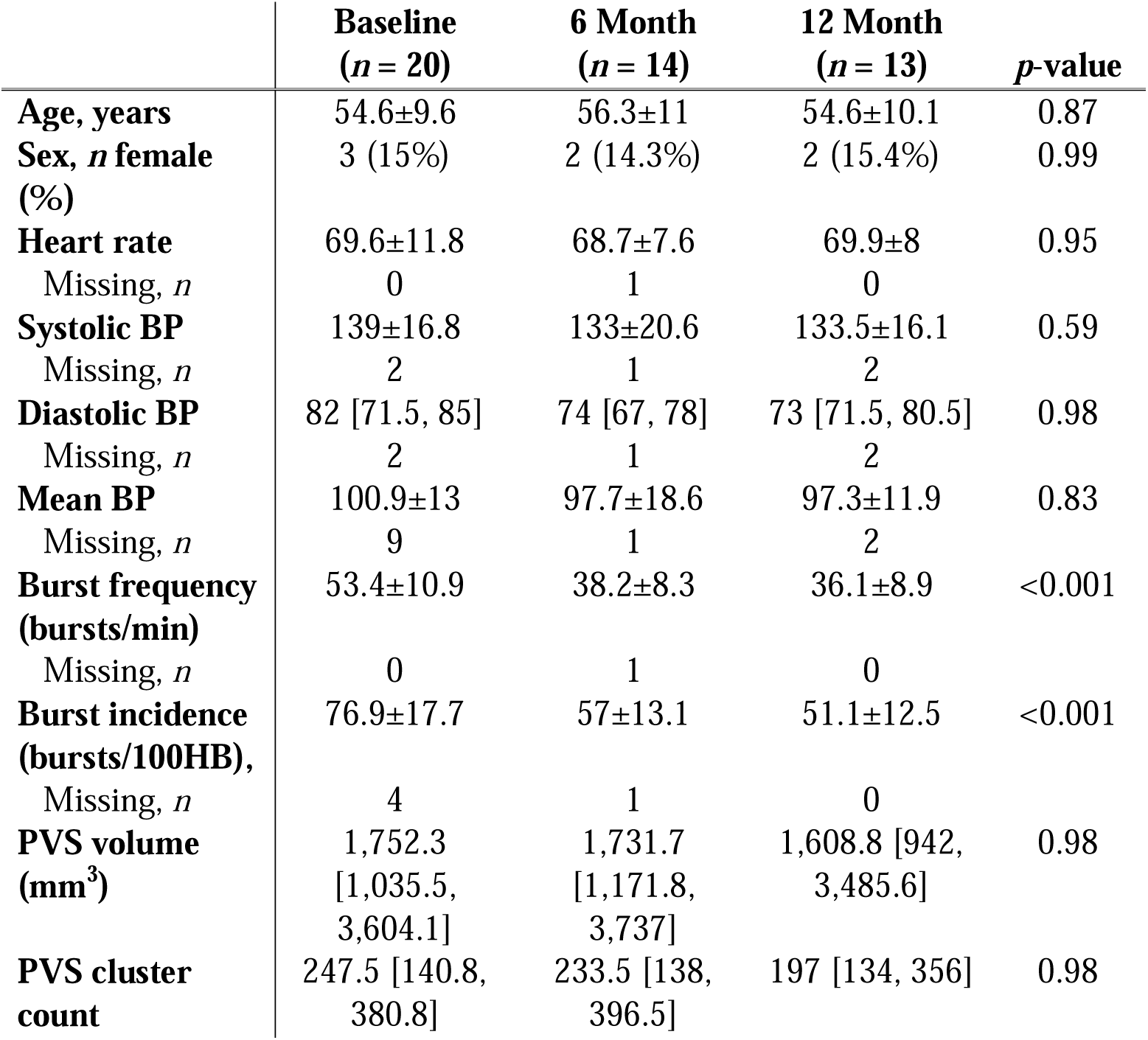
Clinical data and physiological characteristics of the OSA patients across timepoints. Numerical variables are presented as mean±SD or median [Q1, Q3] for normally or non-normally distributed variables, respectively. Within-group differences in categorical variables across timepoints were assessed with chi-squared tests. Within-group differences in numerical variables were compared with Kruskal-Wallis tests.

### Validation of automated perivascular space quantification

Perivascular space cluster counts in representative axial slices were compared between model predictions and manual ratings for the CS and BG. In the CS, there was strong correlation and agreement between methods (Spearman’s ρ=0.88, *p*<0.001; CCC=0.95, 95%CI [0.93, 0.96]). In contrast, the BG showed only moderate correlation and poor agreement (Spearman’s ρ=0.44, *p*<0.001; CCC=0.15, 95%CI [0.1, 0.2]). Agreement between automated and manual counts was also assessed in the midbrain and hippocampus, using the full regions rather than representative slides. Here, PVS measurements demonstrated moderate consistency for both the midbrain (Spearman’s ρ=0.62, *p*<0.001; CCC=0.62, 95%CI [0.39, 0.78]) and hippocampus (Spearman’s ρ=0.78, *p*<0.001; CCC=0.68, 95%CI [0.51, 0.79]) regions. Overall, the consistency of automated versus manual PVS counts was comparable across regions in both control and OSA groups (Table 3).

**Table 3.** Validation of automated perivascular space measurements across brain regions. Spearman’s correlation coefficients (ρ), Lin’s concordance correlation coefficients (CCC), and 95% CI between automated and manual PVS counts across regions of interest (ROI) for the entire cohort, controls, and patients with obstructive sleep apnoea (OSA). Strong agreement was observed for PVS measurements in the centrum semiovale (CS). Moderate agreement was observed for PVS measurements in the midbrain (MB) and hippocampus (HP). Weak agreement was observed for PVS measurements in the basal ganglia (BG).

| Group | ROI | $\rho$ | *p*-value | CCC | 95%CI |
| --- | --- | --- | --- | --- | --- |
| **All** | CS | 0.88 | <0.001 | 0.95 | 0.93–0.96 |
|  | BG | 0.44 | <0.001 | 0.15 | 0.1–0.2 |
|  | MB | 0.62 | <0.001 | 0.62 | 0.39–0.78 |
|  | HP | 0.78 | <0.001 | 0.68 | 0.51–0.79 |
| **Control** | CS | 0.86 | <0.001 | 0.94 | 0.92–0.96 |
|  | BG | 0.36 | 0.001 | 0.18 | 0.09–0.27 |
|  | MB | 0.68 | <0.001 | 0.69 | 0.41–0.86 |
|  | HP | 0.71 | <0.001 | 0.54 | 0.25–0.74 |
| **OSA** | CS | 0.9 | <0.001 | 0.95 | 0.92–0.97 |
|  | BG | 0.61 | <0.001 | 0.13 | 0.08–0.19 |
|  | MB | 0.64 | 0.002 | 0.57 | 0.2–0.79 |
|  | HP | 0.84 | <0.001 | 0.77 | 0.56–0.89 |

### The effect of OSA and CPAP therapy on perivascular spaces

We used GLMs to assess group differences in baseline PVS metrics, adjusting for age, sex, and TBV (Figures 1-2). Compared to controls at baseline, PVS volume was significantly higher in the OSA group at baseline (exp(β)=1.65, 95%CI [1.07, 2.51], *p*=0.012, *p*_FDR_=0.057) and 6-month post-CPAP (exp(β)=1.69, 95%CI [1.07, 2.55], *p*=0.015, *p*_FDR_=0.057), but not at 12-month post-CPAP (exp(β)=1.56, 95%CI [1.03, 2.39], *p*=0.054, *p*_FDR_=0.13). Similarly, whole-brain PVS cluster counts were significantly higher in the OSA group than controls at baseline (exp(β)=1.51, 95%CI [1.1, 2.04], *p*=0.011, *p*_FDR_=0.057) and 6-months post-CPAP (exp(β)=1.55, 95%CI [1.11, 2.12], *p*=0.013, *p*_FDR_=0.057) but not 12-months post-CPAP (exp(β)=1.39, 95%CI [0.97, 1.92], *p*=0.072, *p*_FDR_=0.15).

**Figure 1.**
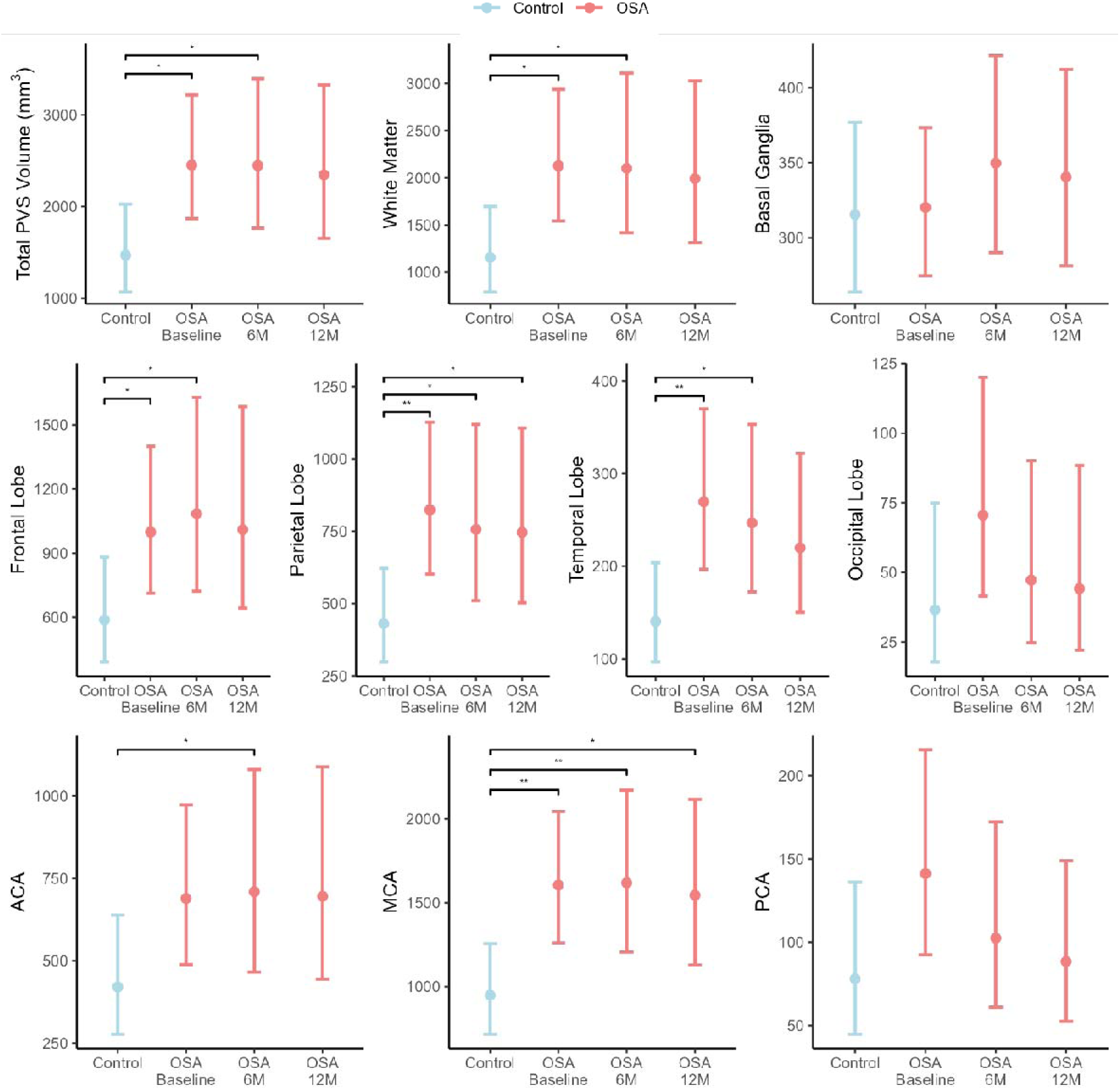
Perivascular space volumes across brain regions in OSA patients and controls. Line plots show estimated marginal means (±95% CI) from generalised linear models. Blue lines represent controls; red lines represent OSA patients. Separate GLMs were fitted to compared PVS quantities between controls at baseline to OSA at baseline, 6-months, and 12-months post-CPAP. Significant group effects are annotated in each panel. At baseline and 6 months following CPAP therapy, PVS volumes were significantly greater in OSA patients compared with controls across several regions, including the whole brain, white matter, frontal, parietal, and temporal lobes, as well as within the MCA territory. After correction for multiple comparisons, significantly greater PVS volumes in OSA patients remained only in the parietal lobe and MCA territory at baseline, and in the MCA territory at 6 months post-CPAP. At the 12-month timepoint, PVS volumes were significantly larger in OSA patients than controls in the parietal lobe and MCA territory. However, these associations did not remain statistically significant after multiple comparisons correction (*p*_FDR_>0.05). Significance: \**p* < 0.05, \*\**p* < 0.01, \*\*\**p* < 0.001.

**Figure 2.**
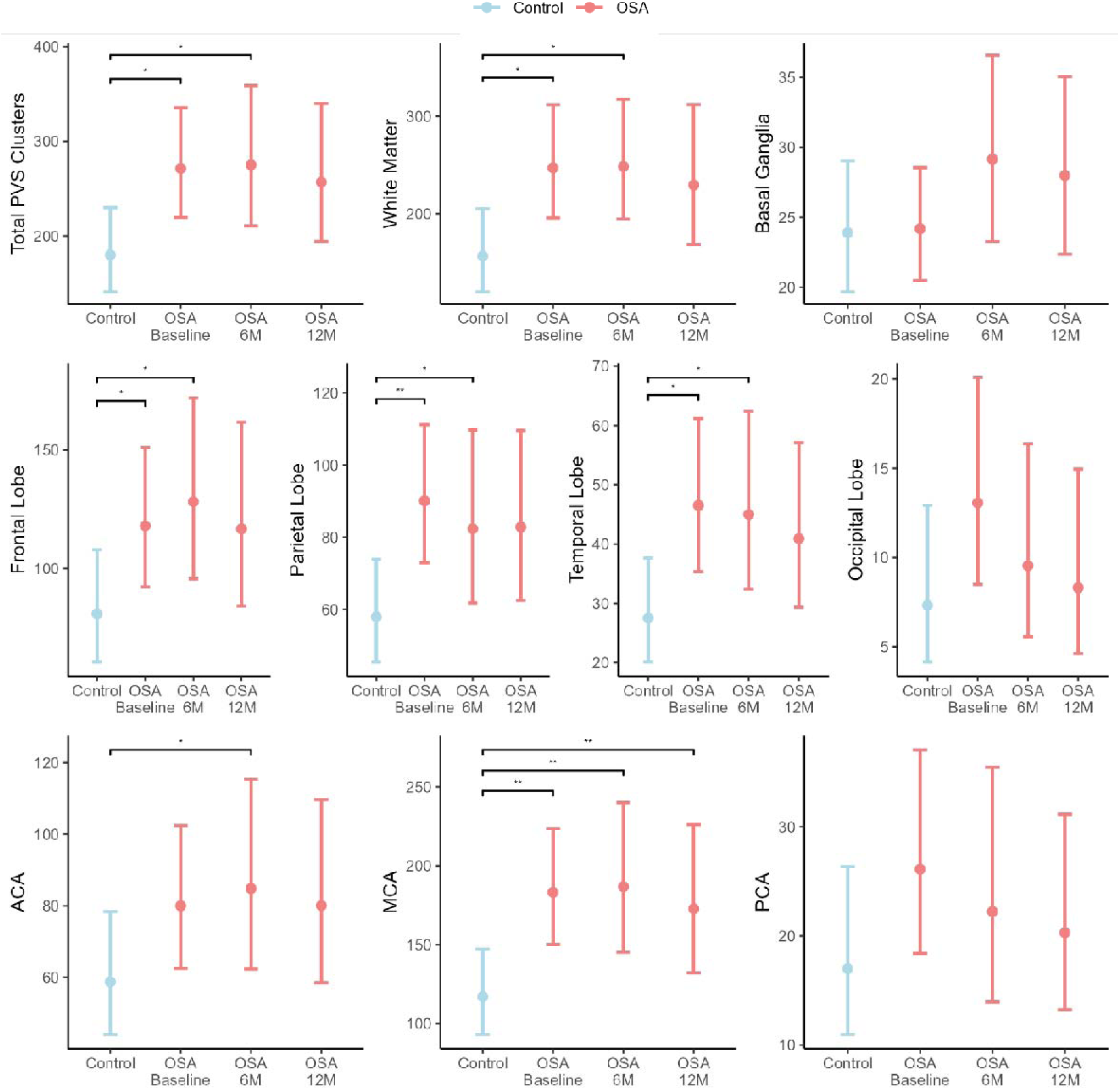
Perivascular space cluster counts across brain regions in OSA patients and controls. Line plots show estimated marginal means (±95% CI) from generalised linear models. Blue lines represent controls; red lines represent OSA patients. Separate GLMs were fitted to compared PVS quantities between controls at baseline to OSA at baseline, 6-months, and 12-months post-CPAP. Significant group effects are annotated in each panel. At the baseline and 6-month timepoints, PVS cluster counts were significantly greater in OSA patients compared with controls across several regions, including the whole brain, white matter, frontal, parietal, and temporal lobes, as well as the MCA territory. After correction for multiple comparisons, significant associations between OSA and increased PVS cluster counts persisted in the parietal lobe and MCA territory at baseline, and in the white matter and MCA territory at 6 months (*p*_FDR_<0.05). At the 12-month timepoint, PVS cluster counts in the MCA territory were significantly greater in OSA patients compared with controls; however, this association did not remain statistically significant after multiple comparisons correction (*p*_FDR_>0.05). Significance: \**p* < 0.05, \*\**p* < 0.01, \*\*\**p* < 0.001.

In the white matter, a similar pattern of results emerged, with WM-PVS volume and WM-PVS cluster counts being significantly higher in OSA at baseline (volume: exp(β)=1.8, 95%CI [1.14, 2.81], *p*=0.013, *p*_FDR_=0.057; count: exp(β)=1.55, 95%CI [1.11, 2.14], *p*=0.011, *p*_FDR_=0.057) and 6-months post-CPAP (volume: exp(β)=1.88, 95%CI [1.1, 3.02], *p*=0.014, *p*_FDR_=0.057; counts: exp(β)=1.67, 95%CI [1.19, 2.41], *p*=0.004, *p*_FDR_=0.041) but not after 12 months of CPAP (volume: exp(β)=1.69, 95%CI [1, 2.81], *p*=0.057, *p*_FDR_=0.13; counts: exp(β)=1.42, 95%CI [0.96, 2.06], *p*=0.081, *p*_FDR_=0.15), compared to controls at baseline. After adjusting for confounds, neither BG-PVS volumes nor cluster counts were associated with OSA diagnosis at any timepoint (*p*>0.05).

Furthermore, we examined PVS metrics across the four lobes of the cerebral cortex. At baseline, OSA patients showed significantly greater PVS volumes and counts in the frontal (volume: exp(β)=1.89, 95%CI [1.14, 3.25], *p*=0.019, *p*_FDR_=0.10; counts: exp(β)=1.61, 95%CI [1.08, 2.33], *p*=0.015, *p*_FDR_=0.099), parietal (volume: exp(β)=1.84, 95%CI [1.12, 3.03], *p*=0.017, *p*_FDR_=0.041; counts: exp(β)=1.49, 95%CI [1.01, 2.18], *p*=0.037, *p*_FDR_=0.041) and temporal lobes (volume: exp(β)=1.81, 95%CI [1.1, 2.88], *p*=0.012, *p*_FDR_=0.057; counts: exp(β)=1.67, 95%CI [1.09, 2.57], *p*=0.016, *p*_FDR_=0.057), but not the occipital lobe (Supplementary Table 2).

After 6 months of CPAP therapy, PVS measures across these three lobar regions remained significantly elevated in OSA patients than in controls (all *p*<0.05; Supplementary Table 2); however, these associations did not remain statistically significant following multiple comparisons correction (*p*_FDR_>0.05).

In contrast, after 12 months of CPAP, no significant differences in PVS volumes or cluster counts were observed between OSA patients and controls in the frontal (volume: exp(β)=1.68, 95%CI [0.94, 2.87], *p*=0.085; counts: exp(β)=1.4, 95%CI [0.92, 2.05], *p*=0.12) or temporal lobes (volume: exp(β)=1.54, 95%CI [0.94, 2.38], *p*=0.085; counts: exp(β)=1.44, 95%CI [0.95, 2.12], *p*=0.089) lobes. In the parietal lobe, only PVS volume remained significantly greater in OSA patients (exp(β)=1.71, 95%CI [1.02, 2.77], *p*=0.036, *p*_FDR_=0.099), whereas cluster counts were not significantly different between groups (exp(β)=1.37, 95%CI [0.96, 1.92], *p*=0.081, *p*_FDR_=0.15), Across the vascular territories, PVS metrics within the MCA territory were significantly greater in OSA patients compared to controls at baseline (volume: exp(β)=1.68, 95%CI [1.19, 2.36], *p*=0.003, *p*_FDR_=0.041; counts: exp(β)=1.56, 95%CI [1.18, 2.12], *p*=0.003, *p*_FDR_=0.041) and at 6 months (volume: exp(β)=1.71, 95%CI [1.16, 2.45], *p*=0.005, *p*_FDR_=0.045; counts: exp(β)=1.62, 95%CI [1.19, 2.18], *p*=0.003, *p*_FDR_=0.041). At 12 months, PVS enlargement in the MCA territory remained evident (volume: exp(β)=1.71, 95%CI [1.02, 2.77], *p*=0.024, *p*_FDR_=0.075; counts: exp(β)=1.44, 95%CI [1.04, 1.96], *p*=0.036, *p*_FDR_=0.099), although these associations did not remain statistically significant after multiple comparisons correction (*p*_FDR_>0.05).

No group differences were observed in the ACA or PCA territories at any timepoint (all *p*>0.05), with the exception of PVS measurements in the ACA territory at the 6 months (volume: exp(β)=1.76, 95%CI [1.02, 3.13], *p*=0.045, *p*_FDR_=0.11; counts: exp(β)=1.49, 95%CI [0.99, 2.2], *p*=0.049, *p*_FDR_=0.12).

Across the midbrain and hippocampal regions, PVS cluster counts in the hippocampus were significantly greater in OSA patients than in controls at the 6-month timepoint (exp(β)=2.13, 95%CI [0.4, 2.31], *p*=0.003, *p*_FDR_=0.041). No other PVS metric in these regions showed significant group differences across the three timepoints (all *p*>0.05).

### Associations with physiological parameters

Generalised linear models were refitted to assess associations between PVS metrics and physiological parameters in OSA participants. After adjusting for confounders, mean blood pressure was positively associated with PVS volume in the parietal lobe (exp(β)=1.02, 95%CI [1, 1.05], *p*=0.044) and MCA territory (exp(β)=1.02, 95%CI [1, 1.04], *p*=0.042). Heart rate was inversely associated with PVS cluster counts in the temporal lobe (exp(β)=0.97, 95%CI [0.95, 0.98], *p*=0.014) and hippocampus (exp(β)=0.97, 95%CI [0.92, 0.99], *p*=0.017).

No significant associations were observed between PVS metrics and systolic blood pressure, MSNA burst rate, or burst incidence (all *p*>0.05). Likewise, no additional associations were found when analyses were extended to whole-brain, white matter, basal ganglia, lobar, vascular territory, midbrain, or hippocampal PVS metrics (all *p*>0.05). However, none of the observed associations remained statistically significant (*p*_FDR_>0.05) after correcting for multiple comparisons.

## Discussion

In this study, we observed region-specific changes in MRI-visible perivascular spaces associated with OSA. PVS burden was significantly greater in OSA patients compared to controls, particularly in the white matter, frontal, parietal and temporal lobes, as well as the middle cerebral artery territory. Following 12 months of CPAP therapy, PVS measurements in the frontal and temporal lobes were no longer significantly higher in OSA patients. However, PVS measurements in the parietal lobe and MCA territory remained significantly elevated post-CPAP compared to controls.

Our findings support the growing body of evidence linking OSA to widespread neuroanatomical and functional alterations. Previous studies have reported a connection between OSA and a range of neuroanatomical changes, including white matter lesions (Ho et al., 2018), cortical thinning across the temporal, frontal, and parietal lobes (Joo et al., 2013; Li et al., 2023), and compromised white matter integrity in the basal ganglia and limbic regions (Rostampour et al., 2020). In addition to these structural changes, alterations in MRI-visible perivascular spaces have also been observed in OSA.

Traditional methods of MRI-PVS assessment involve the counting of PVS clusters in a subset of axial slices per region of interest (Paradise et al., 2020; Potter et al., 2015). For example, higher PVS enlargement scores, defined using rating scales, in the centrum semiovale have been positively correlated with more severe apnoea-hypopnea indices in a cohort of OSA patients (W.-J. Lee et al., 2021). However, a recent meta-analysis reported that the evidence of PVS enlargement in OSA remains inconclusive (G. Lee et al., 2023).

Consistent with previous studies (Jia et al., 2021; W.-J. Lee et al., 2021), we observed significantly enlarged PVS in the white matter of OSA patients compared to controls. Furthermore, by parcellating PVS by lobar regions, we found that enlargement was localised to the frontal, parietal, and temporal lobes. During healthy adolescence, a higher prevalence of PVS is observable in the temporal and parietal lobes compared to the frontal and occipital lobes (Piantino et al., 2020). Our observation could therefore reflect mechanisms that amplify the regional susceptibility to PVS enlargement, especially in the parietal lobe, such as heightened vascular pulsatility.

Vascular territory-based parcellation of PVS revealed significant enlargement in the ACA and MCA regions among OSA patients. While the ACA territory supplies the medial aspects of the frontal and parietal lobes, the MCA branches primarily perfuse their lateral aspects. It is therefore plausible that OSA-related alterations in these lobar regions are facilitated by disruptions in vascular dynamics along the arterial networks of the ACA and MCA.

Importantly, our results extend previous studies by assessing the effects of CPAP therapy on PVS measures over a 12-month period. We found that regional PVS changes associated with OSA are reversible following 12 months of CPAP therapy. Reductions in PVS magnitudes appear to occur gradually. PVS measurements remained significantly enlarged in OSA patients after 6 months of CPAP compared to controls, whereas after 12 months of treatment, PVS metrics in most brain regions were no longer significantly different from those observed in controls. This likely suggests that PVS alterations are not primarily driven by acute effects, such as hypertension, which can improve after a single night of CPAP (Logan et al., 2003), but rather by more gradual structural changes, such as vascular remodeling, or a combination of structural and dynamic functional processes. Together, our findings suggest that long-term CPAP therapy leads to regional reductions in PVS burden and may contribute to the restoration of normal perivascular fluid dynamics.

The basal ganglia are another key region where PVS enlargement is commonly studied. Using rating scales, enlarged PVS in the basal ganglia has been linked to poor sleep efficiency (Del Brutto et al., 2019) and OSA diagnosis (Huang et al., 2019). Recent studies using voxel-wise quantification of PVS have led to links between BG-PVS enlargement and poor sleep quality (Shih et al., 2023), interrupted sleep being uncovered (Aribisala et al., 2020). By contrast, we found no significant link between OSA and BG-PVS enlargement.

Hypertension is a well-established risk factor for basal ganglia PVS enlargement (Francis et al., 2019). Increased vascular pulsatility promotes fluid influx into the perivascular spaces of the basal ganglia (Wardlaw et al., 2020). Moreover, there is evidence that the elevated MSNA in hypertension is associated with basal ganglia (Rim et al., 2024). Additionally, OSA frequently co-occurs with hypertension (Lévy et al., 2015). In our previous studies, we showed that OSA is associated with altered neuronal activity in key autonomic brainstem nuclei, including the dorsolateral pons and rostral ventrolateral medulla, which play a central role in regulating systemic vascular tone and blood pressure (Fatouleh et al., 2014; Lundblad et al., 2014, 2015). Thus, there is a strong physiological basis supporting the presence of BG-PVS enlargement in OSA. This null finding may stem from the segmentation model’s limited sensitivity to PVS in the basal ganglia. Alternatively, our modest sample size may have lacked sufficient statistical power to identify subtle alterations in basal ganglia PVS. Insufficient statistical power may also have contributed to the absence of significant associations between PVS measures and physiological variables, such as systolic blood pressure and muscle sympathetic nerve activity.

In OSA, several structural and functional changes have been documented in the brainstem. For example, voxel-based morphometry studies have reported increased midbrain grey mater volumes in untreated OSA (Lundblad et al., 2014), and altered functional connectivity and diffusivity within brainstem nuclei have also been noted (Lundblad et al., 2014, 2015). However, we found no significant differences in midbrain PVS between OSA patients and controls, nor were these measures significantly altered after CPAP treatment. Similarly, we did not find significant differences in hippocampal PVS burden between OSA patients and controls. Given that MRI-PVS visibility is limited in these small brain regions, a larger sample size or greater imaging resolution than our 3T MRI may be required to detect subtle OSA-related changes in these regions.

### Limitations

Several limitations of our study should be acknowledged. First, our study relied solely on structural MRI to characterise perivascular spaces, limiting our ability to investigate the underlying causal mechanisms. OSA is associated with a wide range of physiological, vascular, and neuronal changes. The primary mechanism responsible for OSA-related PVS enlargement, and its reversal with CPAP, remains to be determined. Whether normalisation of PVS morphology affects glymphatic clearance and neuronal homeostasis also remains unknown.

Future studies incorporating multimodal imaging will be essential to clarify the biological processes involved in OSA-related perivascular space changes. For example, functional MRI could be used to explore whether PVS changes are linked with alterations in functional connectivity or regional cerebral blood flow. Diffusion MRI may help determine whether changes in fluid diffusivity and white matter integrity are linked to PVS enlargement.

Finally, the modest sample size and partial attrition at follow-up timepoints may have limited the statistical power to detect subtle PVS changes, particularly in the smaller brain regions. Future studies with larger cohorts, more frequent imaging timepoints, and cognitive assessments will be essential to validate and expand upon our findings.

## Conclusion

Obstructive sleep apnoea induces region-specific enlargement of perivascular spaces, with notable expansion in the frontal lobe and middle cerebral artery territory. Importantly, these alterations were reversible with effective CPAP therapy. After 12 months of treatment, PVS burden in these regions were no longer elevated compared to healthy controls. Our findings suggest that CPAP therapy can reverse alterations in perivascular spaces within several brain regions, possibly restoring normal glymphatic function. Future longitudinal studies with larger cohorts are needed to fully elucidate the long-term impact of OSA on PVS integrity.

## Data Availability

The dataset analysed in the current study are available from the corresponding author upon reasonable request.

## Acknowledgements

All MRI scanning was conducted at Neuroscience Research Australia.

## Funding

This work was supported by National Health and Medical Research Council of Australia Grant 1007557.

## Notes

**Support:** This research was supported by an Australian Government Research Training Program (RTP) Scholarship.

### Competing Interest Statement

The authors have declared no competing interest.

