## Supplementary Table for "Longitudinal effects of CPAP therapy on MRI-Perivascular spaces in obstructive sleep apnoea"

**Supplementary Table 1.** Total and regional perivascular space (PVS) metrics stratified by group. PVS volume (mm^3^) and cluster counts are shown for the total brain, white matter (WM), basal ganglia (BG), white matter lobes, arterial vascular territories, midbrain (MB), and hippocampus (HP). Values are presented as median [Q1, Q3]. WM = white matter; BG = basal ganglia; ACA = anterior cerebral artery; MCA = middle cerebral artery; PCA = posterior cerebral artery; MB = midbrain; HP = hippocampus.

|  | **Controls (*n*=20)** | **OSA (*n*=20)** | **Total (*N*=40)** | ***p*-value** |
| --- | --- | --- | --- | --- |
| **PVS volume (mm^3^)** | 1,195.1 [945.3, 1,493.4] | 1,752.3 [1,035.5, 3,604.1] | 1,433.1 [988.3, 2,065.2] | 0.033 |
| **PVS cluster count** | 155.5 [119.3, 205.3] | 247.5 [140.8, 380.8] | 165 [123.5, 267.5] | 0.017 |
| **WM PVS volume (mm^3^)** | 917.5 [562.6, 1,211.2] | 1,349.8 [740.5, 3,294.2] | 1,032.2 [636.2, 1,903.9] | 0.032 |
| **WM PVS clusters** | 132 [88, 176.8] | 200.5 [118.5, 355.8] | 151.5 [106.3, 255] | 0.018 |
| **BG PVS volume (mm^3^)** | 296.6 [261.4, 359.4] | 321.6 [247, 430.5] | 300.6 [261.4, 419.6] | 0.575 |
| **BG PVS clusters** | 20.5 [17, 27.3] | 24.5 [15.5, 32.5] | 21 [16.8, 28.3] | 0.232 |
| **Frontal lobe PVS volume (mm^3^)** | 454.6 [247.8, 630.7] | 784.7 [393, 1564.7] | 535.9 [252.2, 938.1] | 0.059 |
| **Frontal lobe PVS clusters** | 67.5 [45.8, 91] | 118 [60.5, 165.5] | 75.5 [46, 128.5] | 0.036 |
| **Parietal lobe PVS volume (mm^3^)** | 391 [194.2, 448.4] | 576.2 [284.4, 1,180.3] | 408.4 [251.9, 726.4] | 0.015 |
| **Parietal lobe PVS clusters** | 55.5 [36, 62.5] | 75 [50.8, 127] | 58 [45.5, 94.5] | 0.007 |
| **Temporal lobe PVS volume (mm^3^)** | 103.6 [75.7, 151.8] | 190.8 [102.4, 341.2] | 120 [75.7, 220.9] | 0.033 |
| **Temporal lobe PVS clusters** | 20 [16, 35.8] | 36.5 [15.8, 60.5] | 25.5 [15.8, 41.5] | 0.04 |
| **Occipital lobe PVS volume (mm^3^)** | 15.7 [3.6, 36.7] | 27.2 [4.9, 65.6] | 17 [3.9, 42.6] | 0.147 |
| **Occipital lobe PVS clusters** | 5 [1.8, 6.8] | 8 [2, 14.8] | 5 [2, 10] | 0.109 |
| **ACA PVS volume (mm^3^)** | 344.2 [185.9, 507.6] | 412.4 [254.4, 1142.3] | 388.4 [199.9, 665.4] | 0.073 |
| **ACA PVS clusters** | 54.5 [36, 68] | 56.5 [44.8, 111.3] | 56.5 [37.8, 85.5] | 0.071 |
| **MCA PVS volume (mm^3^)** | 765.7 [621, 976] | 1,264.3 [724.9, 2,151.7] | 937.8 [650.2, 1,484.7] | 0.02 |
| **MCA PVS clusters** | 99.5 [74, 127] | 171.5 [97.5, 244.3] | 109.5 [82.8, 186.5] | 0.009 |
| **PCA PVS volume (mm^3^)** | 59.7 [27.2, 74.6] | 62.9 [23.3, 140.1] | 59.7 [24.1, 96.4] | 0.121 |
| **PCA PVS clusters** | 13.5 [7.5, 17.3] | 15.5 [8.3, 31.3] | 13.5 [7.5, 21] | 0.139 |
| **MB PVS volume (mm^3^)** | 13.1 [9.2, 18.8] | 13.8 [3.8, 23.4] | 13.1 [6.1, 23.4] | 0.403 |
| **MB PVS clusters** | 3.5 [3, 5] | 4.5 [2.8, 6] | 4 [3, 5.3] | 0.333 |
| **HP PVS volume (mm^3^)** | 3.9 [2.1, 11] | 2.6 [0, 12.1] | 3.9 [0.5, 12] | 0.938 |
| **HP PVS clusters** | 2 [1, 3] | 1 [0, 3] | 1.5 [0.8, 3] | 0.877 |

**Supplementary Table 2.** Generalised linear model (GLM) results for total perivascular space (PVS) volume and cluster counts. Exponentiated fixed-effect estimates (β), bootstrapped 95% confidence intervals (CI), *p*-values, and false discovery rate-corrected *p*-values (*p*_FDR_) are shown for the effect of group diagnosis. Models were adjusted for baseline age, sex, and baseline total brain volume (TBV). Separate GLMs were refitted to compare baseline PVS measurements in controls with those of the OSA group at each timepoint (baseline, 6-months [6M] and 12-months [12M] following CPAP therapy). WM: white matter; BG: basal ganglia; ACA: anterior cerebral artery; MCA: middle cerebral artery; PCA: posterior cerebral artery; MB: midbrain; HP: hippocampus.

|  | **Baseline: OSA vs CN** | | | | **6M: OSA vs CN** | | | | **12M: OSA vs CN** | | | |
| --- | --- | --- | --- | --- | --- | --- | --- | --- | --- | --- | --- | --- |
| **Model** | **Exp(β)** | **95%CI** | ***p*-value** | ***p*_FDR_** | **Exp(β)** | **95%CI** | ***p*-value** | ***p*_FDR_** | **Exp(β)** | **95%CI** | ***p*-value** | ***p*_FDR_** |
| **PVS volume** | 1.65 | 1.07–2.51 | 0.01 | 0.057 | 1.69 | 1.07–2.55 | 0.015 | 0.057 | 1.56 | 1.03–2.39 | 0.054 | 0.13 |
| **PVS cluster counts** | 1.51 | 1.1–2.04 | 0.01 | 0.057 | 1.55 | 1.11–2.12 | 0.013 | 0.057 | 1.39 | 0.97–1.92 | 0.072 | 0.15 |
| **WM PVS volume** | 1.8 | 1.14–2.81 | 0.01 | 0.057 | 1.88 | 1.1–3.02 | 0.014 | 0.057 | 1.69 | 1–2.81 | 0.057 | 0.13 |
| **WM PVS cluster counts** | 1.55 | 1.11–2.14 | 0.01 | 0.057 | 1.67 | 1.19–2.41 | 0.004 | 0.041 | 1.42 | 0.96–2.06 | 0.081 | 0.15 |
| **BG PVS volume** | 0.99 | 0.78–1.24 | 0.96 | 0.98 | 1.06 | 0.83–1.35 | 0.63 | 0.74 | 1.07 | 0.84–1.36 | 0.57 | 0.69 |
| **BG PVS cluster counts** | 1.13 | 0.89–1.44 | 0.36 | 0.49 | 1.25 | 0.95–1.63 | 0.15 | 0.23 | 1.23 | 0.93–1.64 | 0.16 | 0.24 |
| **Frontal lobe PVS volume** | 1.66 | 1.04–2.64 | 0.04 | 0.1 | 1.89 | 1.14–3.25 | 0.019 | 0.062 | 1.68 | 0.94–2.87 | 0.085 | 0.15 |
| **Frontal lobe PVS cluster counts** | 1.46 | 1.02–2.08 | 0.04 | 0.099 | 1.61 | 1.08–2.33 | 0.015 | 0.057 | 1.4 | 0.92–2.05 | 0.12 | 0.18 |
| **Parietal lobe PVS volume** | 1.9 | 1.18–2.93 | <0.001 | 0.041 | 1.84 | 1.12–3.03 | 0.017 | 0.058 | 1.71 | 1.02–2.77 | 0.036 | 0.099 |
| **Parietal lobe PVS cluster counts** | 1.56 | 1.16–2.06 | <0.001 | 0.041 | 1.49 | 1.01–2.18 | 0.037 | 0.099 | 1.37 | 0.96–1.92 | 0.081 | 0.15 |
| **Temporal lobe PVS volume** | 1.92 | 1.2–3.03 | 0.01 | 0.057 | 1.81 | 1.1–2.88 | 0.012 | 0.057 | 1.54 | 0.94–2.38 | 0.085 | 0.15 |
| **Temporal lobe PVS cluster counts** | 1.68 | 1.1–2.55 | 0.02 | 0.057 | 1.67 | 1.09–2.57 | 0.016 | 0.058 | 1.44 | 0.95–2.12 | 0.089 | 0.15 |
| **Occipital lobe PVS volume** | 1.9 | 0.88–4.11 | 0.1 | 0.17 | 1.4 | 0.51–3.27 | 0.46 | 0.58 | 1.24 | 0.48–2.88 | 0.66 | 0.74 |
| **Occipital lobe PVS cluster counts** | 1.73 | 0.94–3.34 | 0.09 | 0.15 | 1.4 | 0.64–2.91 | 0.37 | 0.5 | 1.15 | 0.51–2.44 | 0.72 | 0.79 |
| **ACA PVS volume** | 1.6 | 0.98–2.68 | 0.07 | 0.15 | 1.76 | 1.02–3.13 | 0.045 | 0.11 | 1.61 | 0.93–2.8 | 0.11 | 0.18 |
| **ACA PVS cluster counts** | 1.36 | 0.94–1.97 | 0.09 | 0.16 | 1.49 | 0.99–2.2 | 0.049 | 0.12 | 1.32 | 0.87–1.95 | 0.17 | 0.25 |
| **MCA PVS volume** | 1.68 | 1.19–2.36 | <0.001 | 0.041 | 1.71 | 1.16–2.45 | 0.005 | 0.045 | 1.59 | 1.06–2.31 | 0.024 | 0.075 |
| **MCA PVS cluster counts** | 1.56 | 1.18–2.12 | <0.001 | 0.041 | 1.62 | 1.19–2.18 | 0.003 | 0.041 | 1.44 | 1.04–1.96 | 0.036 | 0.099 |
| **PCA PVS volume** | 1.84 | 0.99–3.59 | 0.07 | 0.15 | 1.36 | 0.67–2.69 | 0.40 | 0.53 | 1.08 | 0.52–2.06 | 0.83 | 0.89 |
| **PCA PVS cluster counts** | 1.64 | 1.01–2.88 | 0.07 | 0.15 | 1.29 | 0.65–2.43 | 0.43 | 0.55 | 1.12 | 0.65–1.9 | 0.70 | 0.77 |
| **MB PVS volume** | 0.85 | 1.3–0.54 | 0.54 | 0.67 | 0.95 | 1.31–0.84 | 0.86 | 0.91 | 0.98 | 1.32–0.93 | 0.94 | 0.97 |
| **MB PVS cluster counts** | 1 | 0.91–1.71 | 0.18 | 0.26 | 1.16 | 1.18–2.44 | 0.37 | 0.51 | 1.07 | 1.16–1.6 | 0.64 | 0.74 |
| **HP PVS volume** | 1 | 1.46–0.99 | 0.99 | 0.99 | 0.95 | 1.53–0.89 | 0.91 | 0.95 | 0.79 | 1.57–0.6 | 0.61 | 0.73 |
| **HP PVS cluster counts** | 1.01 | 1.01–3.81 | 0.18 | 0.74 | 2.13 | 1.29–19.09 | 0.003 | 0.041 | 0.69 | 1.46–0.37 | 0.32 | 0.45 |
